# Parental Risk Perception: Influence of Disease Perceived Controllability, Experience and Severity

**DOI:** 10.1101/2023.01.25.23284837

**Authors:** Philippe Sylvestre, Pascal Roland Enok Bonong, Tamara Perez, Olivier Drouin

**Author notes:** **Address correspondence to:** Olivier Drouin, Division of General Pediatrics, CHU Sainte-Justine, 3175 Chemin de la Côte-Sainte Catherine, Suite 7939, Montreal (Quebec) H3T 1C5 CANADA.

## Abstract

Optimistic health bias is widespread in adults, impacting responsiveness to information regarding health risks. Comparatively little is known about parents’ perception of their child’s likelihood of developing disease, despite the frequency with which parents make decisions about their child’s health. We hypothesized that parental optimism about their child’s likelihood of developing disease would be greater for diseases perceived as more controllable and those with which parents do not have experience.

Parents of children <18 years complete an online survey. Primary outcome was participants’ perceived comparative likelihood of their child developing appendicitis, dental caries, head lice, leukemia, food allergies, pneumonia, asthma and obesity. Optimism was defined as the belief that one’s child was less likely than average to develop a given disease. Logistic regression models were used to examine the relationship between parental optimism and four independent variables: perceived disease controllability; knowing someone with the disease; child’s history of the disease; perceived disease severity.

Among 374 participants, the proportion of optimistic parents ranged from 35.3% (appendicitis) to 65.0% (obesity). Perceived controllability increased the odds of parental optimism (adjusted odds ratio [ aOR] range: 1.57 for asthma to 4.48 for head lice). Odds of optimism were lower if participants knew somebody with the disease (aOR range: 0.42 for head lice to 0.81 for leukemia) or if the child had a history of the disease (aOR range: 0.27 for dental caries to 0.47 for head lice).

These findings are important to enhance parental counseling effectiveness regarding child health behaviors.

## Introduction

There is growing recognition of the importance of lifestyle behaviors and treatment adherence for pediatric health. (Palfrey et al., 2005) Modifiable behavioral risk factors in childhood, such as unhealthy eating and excessive screen time, increase risk of noncommunicable diseases over one’s lifespan (Halfon & Hochstein, 2002) and non-adherence to preventive treatments contribute considerably to the burden of chronic diseases such as asthma. (Bauman et al., 2002) Helping parents and children engage in healthier behaviors has become central to the practice of pediatrics. (Winnick et al., 2005) As such, much research has focused on helping pediatricians and families identify elements underlying motivation to change behaviors. Among these, both theory (Rosenstock, 1990) and empirical evidence (de Vries et al., 2012; Ferrer & Klein, 2015; Sheeran et al., 2014) suggest that health-related risk perceptions play a central role in people’s decision to engage in preventive health behaviors. However, when asked about their child’s risk of different diseases, most parents consider their child to be at lower than average risk. (Chadi et al., 2020; Drouin et al., 2019; Wright et al., 2017) This bias in risk judgements has been dubbed *unrealistic optimism*, or *optimistic bias*. (Shepperd et al., 2013; Weinstein, 1980)

Optimistic bias is widespread in adults, across a range of health outcomes, (Cho et al., 2013; Clarke et al., 2000; Kreuter & Strecher, 1995; Sargeant et al., 2010; Weinstein, 1980; Weinstein, 1987; Wolde et al., 2019) including risks of alcohol use disorder, (Weinstein, 1980) various cancers, (Clarke et al., 2000; Weinstein, 1980) influenza, (Cho et al., 2013) acute gastro-intestinal illness, (Sargeant et al., 2010) and lead poisoning. (Wolde et al., 2019) Although having a positive outlook on life may provide some health benefits, (Rasmussen et al., 2009) unrealistic optimism may undermine incentives to engage in health-promoting behaviors. (Dillard et al., 2006; Kreuter & Strecher, 1995) For example, smokers with higher levels of optimism are less likely to plan on, (Dillard et al., 2006) and succeed at quitting smoking. (Kreuter & Strecher, 1995) Optimism can also act as a barrier to counseling: people who display optimistic bias tend to adopt avoidant coping strategies, be more defensive during negative health-related feedback, and be less responsive to risk information aimed at promoting healthy behaviors. (Cho et al., 2013; Fowler & Geers, 2015)

Interestingly, adults are more likely to be optimistic regarding events they perceive as being controllable (e.g., drug addictions, sexually transmitted diseases and obesity). (Harris, 1996; Helweg-Larsen & Shepperd, 2001; Klein & Helweg-Larsen, 2002; Weinstein, 1980; Weinstein, 1987) Conversely, they tend to be less optimistic with regards to conditions they have had previous experience with. (Helweg-Larsen & Shepperd, 2001; Weinstein, 1980; Weinstein, 1987) Another potential moderator of optimism is perceived disease severity, although its correlation with unrealistic optimism has shown inconsistent results. (Helweg-Larsen & Shepperd, 2001; Weinstein, 1980; Weinstein, 1987)

Comparatively little is known about parents’ perception of the likelihood that their child develops disease, an important gap given the frequency and importance of decisions made by parents regarding their child’s health. Some studies have examined parents’ risk perception regarding childhood vaccinations, (Hendrix et al., 2014) teenage alcohol and substance use, (Chadi et al., 2020; Napper et al., 2015) cancer, (Sung et al., 2009) and obesity. (Etelson et al., 2003) This evidence suggests that, like adults’ risk perception about themselves, parent’s *proxy* risk perception may also be influenced by an optimistic bias when considering their child’s likelihood of disease. To date however, no study has analyzed the influence of perceived disease controllability, prior experience, and perceived disease severity on parental risk perception. Better understanding parental optimism and its moderators is crucial to developing effective interventions to improve child health. (Helweg-Larsen & Shepperd, 2001)

As such, this study aimed to (1) determine whether parents display unrealistic optimism regarding their child’s likelihood of developing eight different diseases, and (2) explore the contribution of perceived controllability, prior experience, or perceived disease severity on parents’ risk perception. This study tested the following hypotheses: (1) a majority of parents would believe that their child is at lower risk than the average to develop each of eight selected diseases; (2) parents would be more likely to display optimism about diseases perceived as more controllable and (3) diseases for which they had no prior experience; (4) however, there wouldn’t be a significant association between parental optimism and perceived disease severity.

## Materials and methods

### Design and Population

Parents of children aged 0-18 years old completed an online cross-sectional survey in June 2019 using Amazon MTurk. (Crump et al., 2013) Amazon MTurk is an online crowdsourcing service for web-based tasks, and has been used previously to efficiently gather human subject data, and evaluated for experimental behavioural research. (Crump et al., 2013) For this study, respondents were ≥18 years of age, parents of a child aged <18 years and living in Canada or the United States. Each participant received a small monetary sum for taking the survey. The average completion time was approximately ten minutes. Participants were asked to provide answers to the survey regarding their oldest child who was ≤18 years old. Data were managed using REDCap, (Harris et al., 2009) de-identified and stored on a password-protected server hosted at the CHU Sainte-Justine. This study was approved by the ethics review board of the CHU Sainte-Justine.

### Outcome

Primary outcome was participants’ perception of their child’s likelihood of developing eight different pediatric diseases: appendicitis, dental caries, head lice, leukemia, food allergies, pneumonia, asthma, and obesity. Diseases were selected based on the likelihood that they would be known by a majority of participants, and such that they would cover a range of perceived controllability. Parental perception of their child’s likelihood of developing each disease was evaluated with questions in the following format: “Compared to other children his/her age, how likely is your child to develop [ the disease]?” For leukemia, appendicitis, and asthma, parents had the option of selecting “Not applicable” if their child already had the disease. A time frame of 12 months was specified for some diseases (dental caries, pneumonia, head lice, appendicitis) in order to make them more concrete, and to control for potential moderating effects of perceived event distance. (Helweg-Larsen & Shepperd, 2001) Exact formulation of each question can be found in Appendix 1. Similar to other relevant studies, (Shepperd et al., 2018; Weinstein, 1987) comparative risk judgments were measured using a 5-point Likert scale (“much less likely”, “less likely”, “as likely”, “more likely”, “much more likely”). This direct method of eliciting comparative optimism is in contrast with an indirect method (participants providing separate estimates of their personal risk and of a reference population’s risk). Although the indirect method may provide additional information, (Helweg-Larsen & Shepperd, 2001) the direct method provides valid information and is widely used for its simplicity and ease of understanding. (Shepperd et al., 2013)

### Independent variables

For each disease examined, four independent variables were explored: (1) perceived controllability in preventing one’s child from developing the disease (“no control at all”, “a little control”, “a moderate amount of control”, “a lot of control” and “a great deal of control”); (2) knowledge of someone with the disease (yes vs. no); (3) previous history of the disease in the child (yes vs. no); and (4) parental perception of disease severity (100-point scale from “very minor” to “very severe”). The question concerning previous history of the disease in the participant’s child was not included for appendicitis and asthma, given that these conditions do not recur. The exact wording of the questions used to collect the data relating to all these variables is presented in Appendix 1.

### Other covariates

Covariates measured included participants’ age (continuous) and gender (female vs. male), his/her level of educational achievement (“at most high school graduate”, “some college or technical school”, “college graduate”, “completed a graduate degree”), as well as child’s age (continuous) and gender (female vs. male).

### Statistical analysis

Descriptive statistics were presented using absolute and relative frequencies (%), while that of the continuous variables using median and interquartile range (IQR). For each disease, risk perception at the group level was evaluated using one-sample t-tests, as per the widely used method employed in similar relevant studies. (Sargeant et al., 2010; Weinstein, 1987) First, each comparative risk assessment was attributed a score between -2 and +2 (“much less likely” = -2, “less likely” = -1, “as likely” = 0, etc.). Then, one-sample t-tests were used to test the hypothesis that the mean of comparative risk was different from zero: a mean of zero would indicate no bias while a mean below zero would indicate unrealistic optimism at the group level.

Preliminary results of the descriptive analysis guided transformation of the variables for regression analyses. A small number of participants in some categories of the primary outcome required its transformation into a dichotomous variable for logistic regression analyses: “optimistic” (“less likely” or “much less likely”) and “not optimistic” (“as likely”, “more likely” or “much more likely”). Perceived controllability was also recoded into three modalities for the same reason: “no control at all or a little control”; “a moderate amount of control” and “a lot of control or a great deal of control”. Logistic regression was used to estimate the odds ratios (OR) and 95% confidence intervals (CI) of the associations between the main independent variables and parental optimism. For each of the eight diseases considered, three models were estimated: (1) a univariate logistic model for each of the variables examined; (2) a logistic model containing the outcome and the main independent variables; and (3) a logistic model containing the outcome, the main independent variables and the socio-demographic characteristics of the participants and their child. To overcome the problem of missing data and reduce its potential selection bias, we used multiple imputation by chained equations (White et al., 2011) and sensitivity analyses with inverse probability weighting (IPW). Full details of statistical analyses are in Appendix 2. All statistical analyses were performed with Stata/SE 14.2, and the standard threshold of p<0.05 was considered for statistical significance.

## Results

A total of 374 parent participants completed the survey. Their median age was 37.0 years (IQR 32.0 to 43.0 years), 62.3% were female and 62.4% had a college degree. Participants’ children had a median age of 9.0 years (IQR 5.0 to 14.0) and 44.9% were female (Table 1).

**Table 1.**
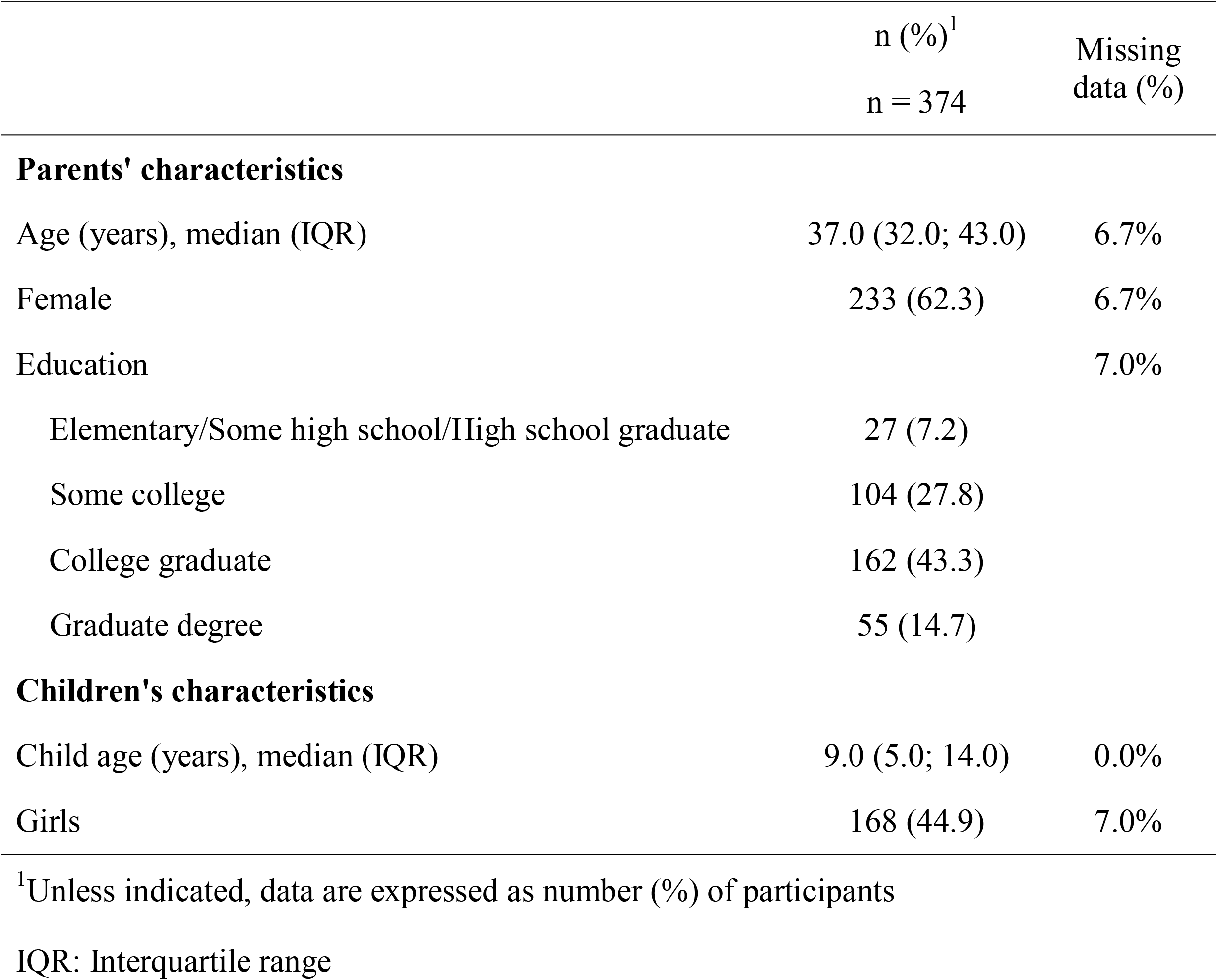
Participants’ socio-demographic data.

For all diseases, there was an optimistic bias at the group level (p<0.001 for all diseases, Table 2). Levels of comparative optimism, defined as parents reporting that their child was “less likely” or “much less likely” than other children to experience a given disease, ranged from 35.3% for appendicitis to 65.0% for obesity. In contrast, the proportion of participants who responded that their child was “more likely” or “much more likely” to develop a given condition ranged from 1.3% for leukemia to 13.9% for dental caries (Figure 1).

**Table 2.**
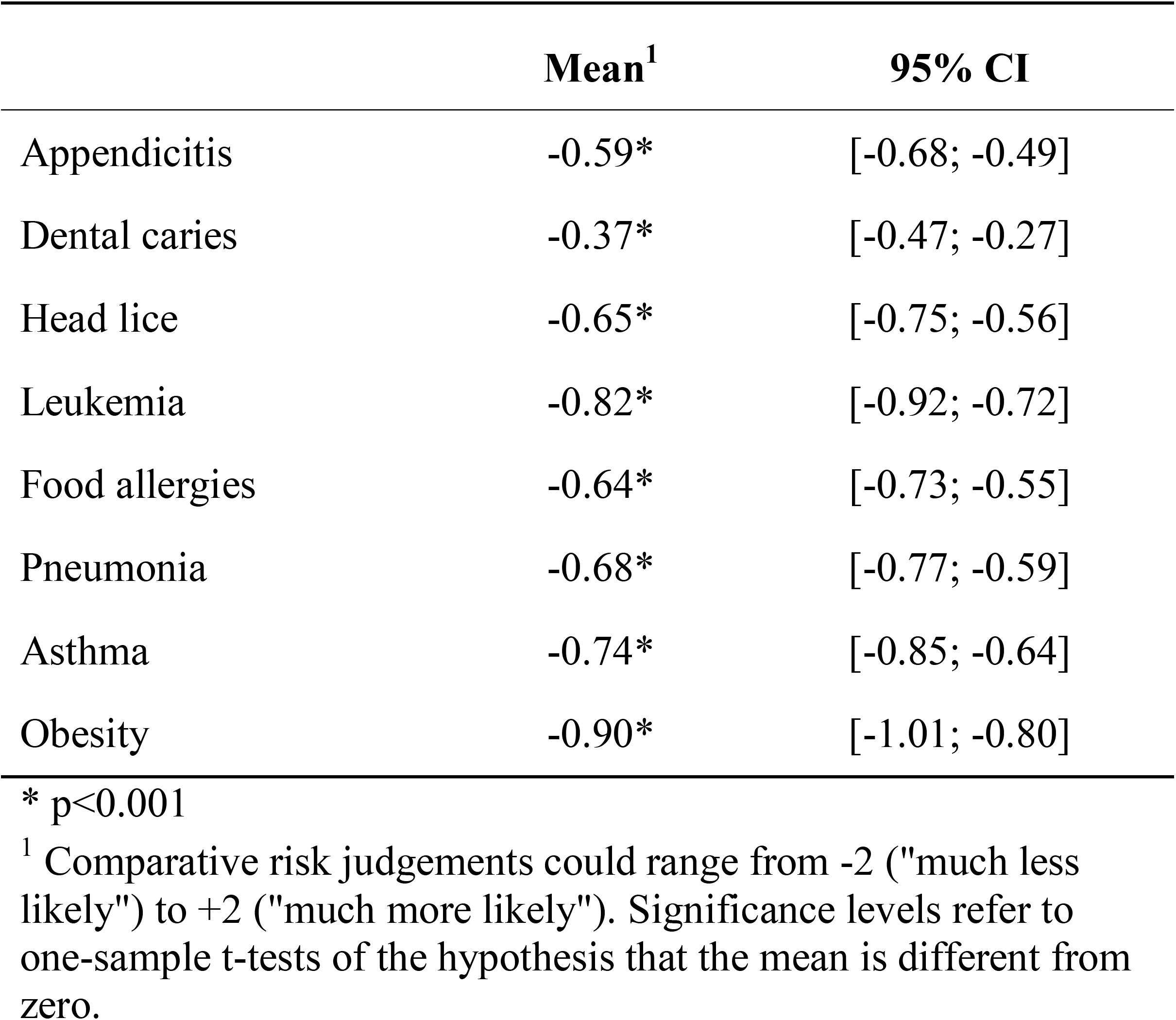
Comparative risk judgements for eight diseases.

**Figure 1:**
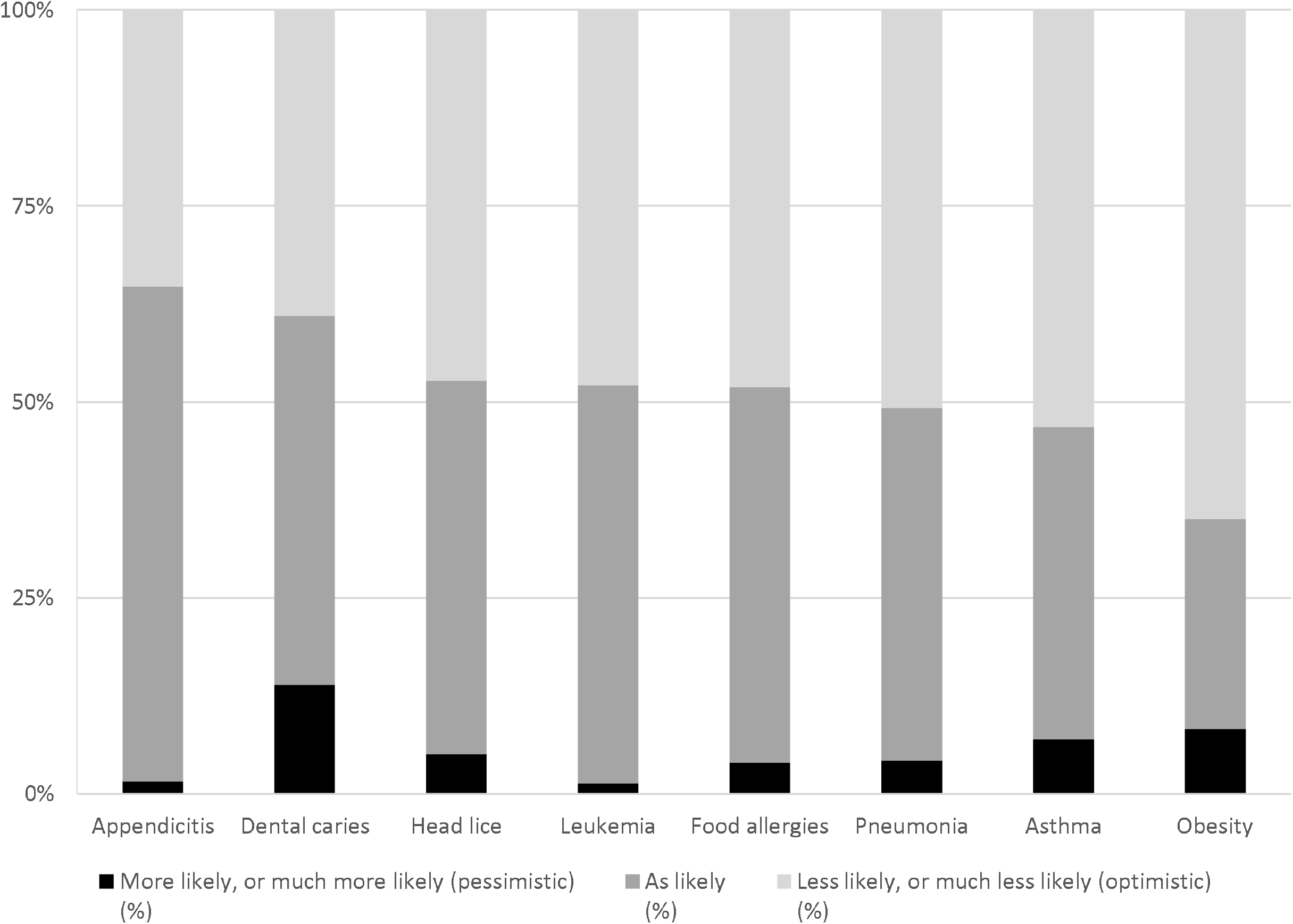
Parents’ perceived likelihood of their child developing eight diseases, compared to the average child.

Participants expressed variable levels of perceived controllability between diseases. While a majority believed they had “no control at all” or “a little control” over their child’s risk of developing appendicitis and leukemia (84.0% and 85.6% respectively), 70.1% felt that they had “a lot of control” or “a great deal of control” in preventing obesity in their child (Figure 2). A majority of participants declared knowing someone else with each of the diseases studied (range 59.4% to 88.2%), with the exception of leukemia (35.6%) (Table 3). While many participants reported that their child had previously had dental caries (43.6%) and hair lice (21.7%), few reported that their child had had any of the other diseases examined (range 1.3% to 12.0%) (Table 3). Finally, perceived severity of each disease varied from a median of 27/100 (IQR 13; 54) for head lice to 99/100 (IQR 95; 100) for leukemia (Table 4).

**Table 3.**
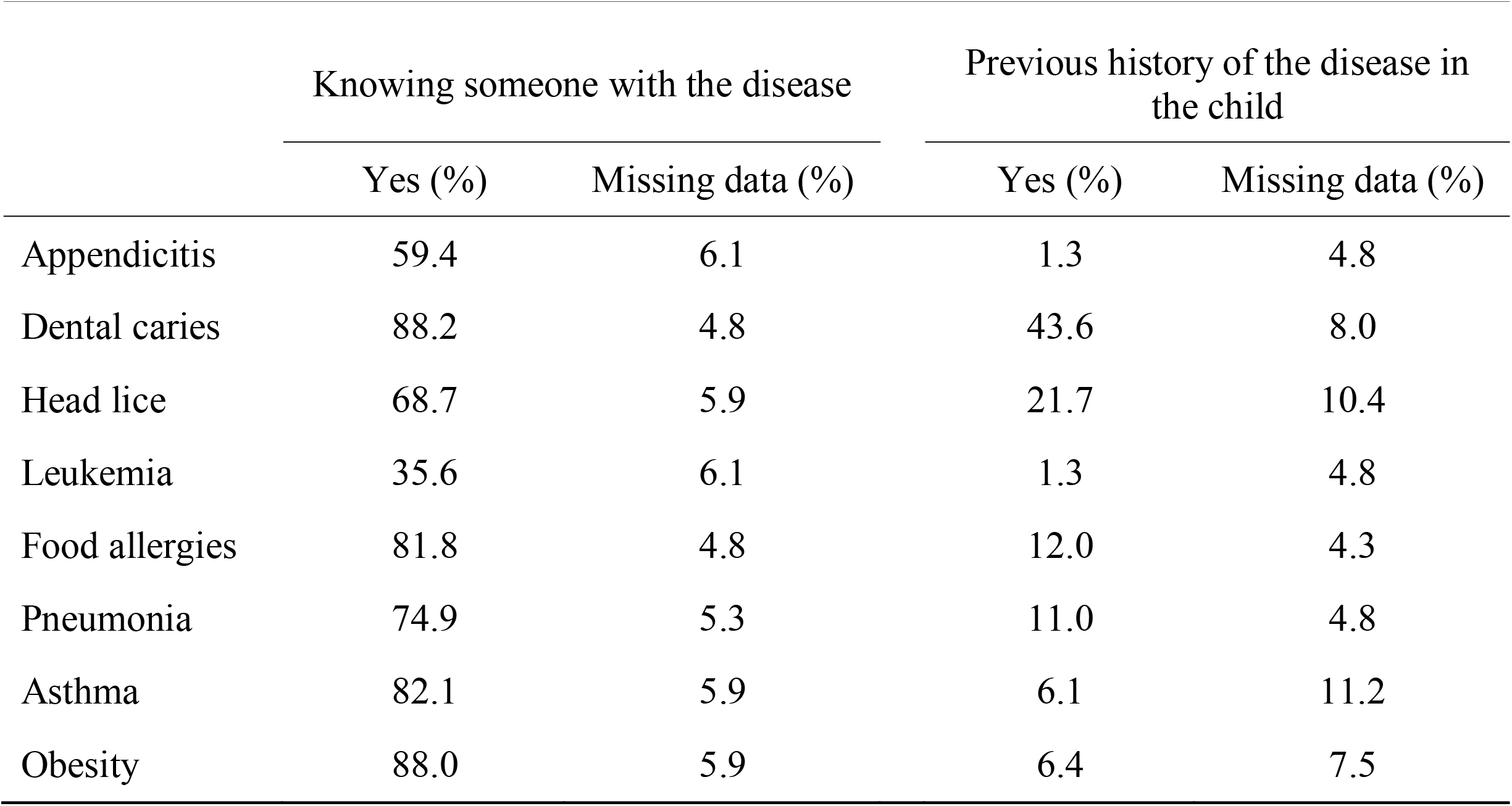
Proportion of parents knowing someone with the disease and proportion of children having previously had the disease.

**Table 4.** Parental perceived severity of eight diseases, on a scale of 0 to 100^1^.

|  | Median | IQR | Missing data (%) |
| --- | --- | --- | --- |
| Appendicitis | 79.0 | 68.0-93.0 | 0.0% |
| Dental caries | 78.0 | 68.0-89.0 | 0.5% |
| Head lice | 80.5 | 64.0-95.0 | 0.5% |
| Leukemia | 71.0 | 53.0-83.0 | 0.3% |
| Food allergies | 74.0 | 60.0-85.0 | 0.0% |
| Pneumonia | 27.0 | 13.0-53.5 | 0.0% |
| Asthma | 99.0 | 95.0-100.0 | 0.0% |
| Obesity | 55.5 | 37.0-70.0 | 0.3% |

<sup>1</sup> 0 considered as very minor, 100 as very severe
IQR: Interquartile range

**Figure 2.**
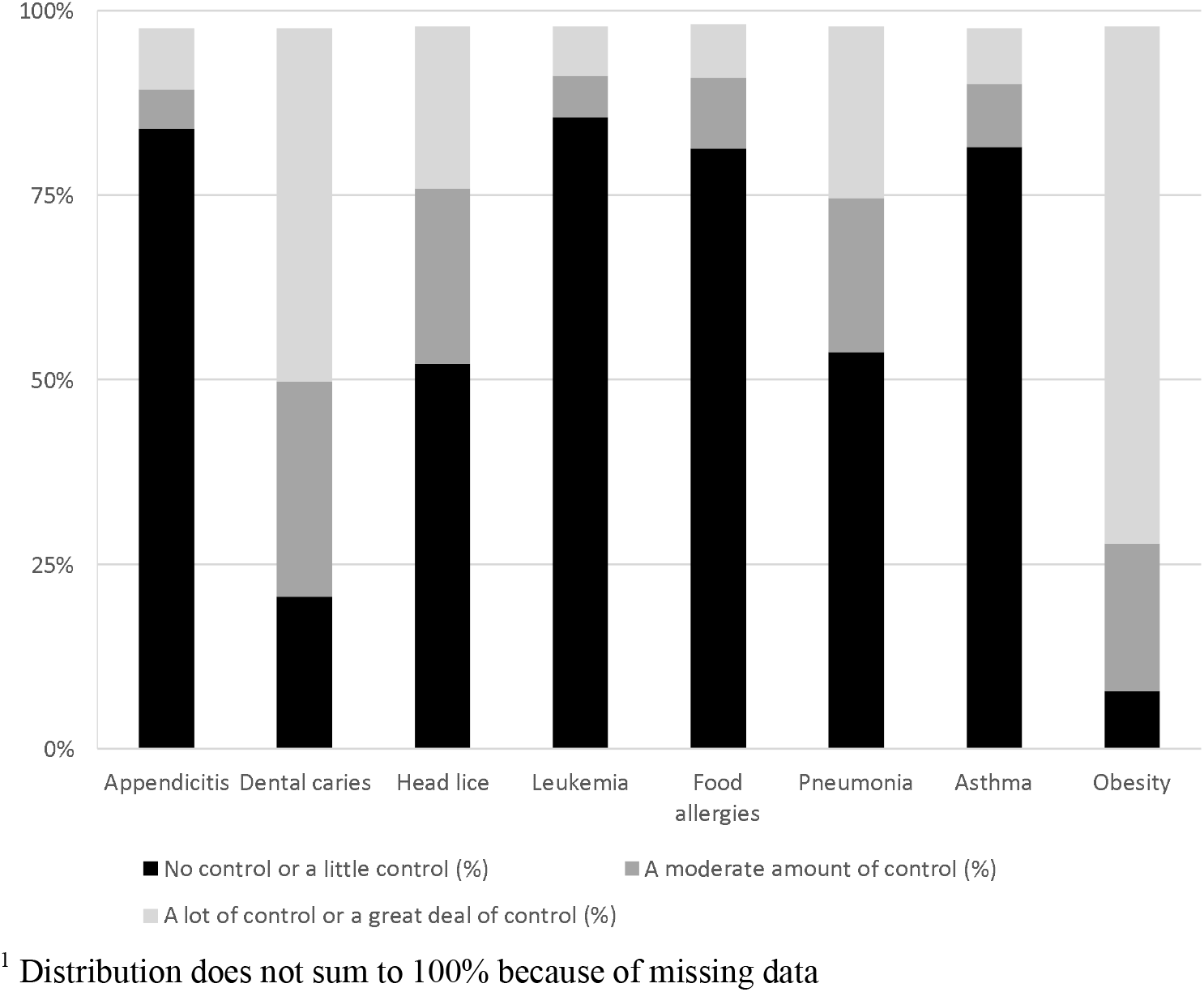
Distribution of participants’ perceived controllability in preventing eight diseases^1^.

For most diseases studied, comparative optimism was correlated with higher perceived controllability (Table 5). When participants perceived a lot of control or a great deal of control, adjusted odds ratios (aOR) of parental optimism ranged from 1.57 to 4.48, which was statistically significant for most diseases (dental caries, pneumonia, leukemia, head lice and obesity). This relationship did not reach statistical significance for asthma, appendicitis, nor food allergies.

**Table 5.** Odds of parental optimism regarding their child’s likelihood of developing eight diseases, depending on parental perceived controllability, experience and perceived disease severity.

| Variables | Adjusted odds ratio for parental optimism<sup>1-3</sup> (95% CI) |  |  |  |  |  |  |  |
| --- | --- | --- | --- | --- | --- | --- | --- | --- |
|  | Appendicitis | Dental caries | Head lice | Leukemia | Food allergies | Pneumonia | Asthma | Obesity |
| *Perceived controllability*<br>(Ref: No control at all or a little control) |  |  |  |  |  |  |  |  |
| A moderate amount of control | **3.07\***<br>(1.15 - 8.18) | 2.04<br>(1.00 - 4.19) | **2.02\***<br>(1.16 - 3.51) | 1.50<br>(0.55 - 4.08) | 0.74<br>(0.34 - 1.61) | 1.42<br>(0.83 - 2.43) | 1.49<br>(0.68 - 3.27) | 1.80<br>(0.69 - 4.66) |
| A lot, or a great deal of control | 1.93<br>(0.85 - 4.39) | **2.64\*\***<br>(1.35 - 5.17) | **4.48\*\***<br>(2.40 - 8.37) | 2.55<br>(1.00 - 6.50) | 2.22<br>(0.92 - 5.36) | **3.57\*\***<br>(2.03 - 6.29) | 1.57<br>(0.65 - 3.77) | **3.04\***<br>(1.29 - 7.18) |
| Knowing someone with the disease (vs. No) | 0.68<br>(0.42 - 1.10) | 0.61<br>(0.25 - 1.51) | **0.42\*\***<br>(0.24 - 0.75) | 0.81<br>(0.52 - 1.28) | **0.48\***<br>(0.25 - 0.94) | 0.77<br>(0.44 - 1.33) | **0.46\***<br>(0.23 - 0.94) | 0.54<br>(0.18 - 1.59) |
| Previous history of the disease in the child (vs. No) | N/A<sup>4</sup> | **0.27\*\***<br>(0.16 - 0.48) | **0.47\***<br>(0.26 - 0.88) | N/A<sup>5</sup> | **0.39\***<br>(0.18 - 0.80) | **0.38\*\***<br>(0.18 - 0.79) | N/A<sup>4</sup> | **0.14\*\***<br>(0.05 - 0.42) |
| Perceived severity of the disease (per 1-point increment) | 1.00<br>(0.99 - 1.01) | 1.00<br>(0.99 - 1.01) | **1.01\***<br>(1.00 - 1.02) | 0.99<br>(0.97 - 1.01) | 0.99<br>(0.98 - 1.00) | 1.00<br>(0.99 - 1.01) | 1.00<br>(0.99 - 1.01) | **1.02\***<br>(1.00 - 1.03) |

\* p<0.05; \*\* p<0.01
1. Parental optimism represents the likelihood that a parent considers their child less or much less likely than average to develop the disease 2. Odds of parental optimism were adjusted for participants' age, gender and level of educational achievement, as well as child's age and gender 3. Results were estimated using some imputation to overcome missing data (0-10% of data for all independent variables, details in Appendix 2) 4. Previous history of the disease in the child was not considered for the analysis of parents' optimism regarding appendicitis and asthma, because these diseases are not diagnosed more than once 5. Previous history of the disease in the child was not considered for the analysis of parents' optimism regarding leukemia because at least 94% of respondents answered "No"

Participants who knew somebody who had been affected by a given disease were less likely to be optimistic regarding their child’s likelihood of developing that disease (Table 5). This relationship was statistically significant for asthma (aOR 0.46; 95% CI 0.23-0.94), head lice (aOR 0.42; 95% CI 0.24-0.75), and food allergies (aOR 0.48; 95% CI 0.25-0.94). Participants were also significantly less likely to be optimistic regarding each of the diseases if their child had previously had the disease (aOR ranging from 0.27 to 0.47) (Table 5). Finally, perceived severity of disease had no statistically significant association with participants’ optimism for any of the disease studied (Table 5).

Participants’ age, gender, and level of education showed no association with parental optimism for most diseases studied (Table 6). Similarly, there was no significant association between the age and gender of the participants’ children and participants’ optimism for the majority of diseases studied. Sensitivity analyses using observed data (without imputation) and weighting of estimates by IPW did not lead to any major difference in the relationships between the dependent and independent variables. Although a few associations gained or lost statistical significance (with a p-value <0.05), the direction and magnitude of effect remained similar between the three analyses (Tables 7a and 7b).

**Table 6.** Odds of parental optimism regarding their child’s likelihood of developing eight diseases, depending on participant’s socio-demographic data.

| Variables | Adjusted odds ratio for parental optimism<sup>1, 2</sup> (95% CI) |  |  |  |  |  |  |  |
| --- | --- | --- | --- | --- | --- | --- | --- | --- |
|  | Appendicitis | Dental caries | Head lice | Leukemia | Food allergies | Pneumonia | Asthma | Obesity |
| Age of the child in years (linear) | 1.03<br>(0.97 - 1.10) | 1.02<br>(0.96 - 1.09) | **1.15\*\***<br>**(1.08 - 1.23)** | 1.01<br>(0.96 - 1.07) | 1.05<br>(1.00 - 1.12) | 1.03<br>(0.97 - 1.09) | 1.03<br>(0.97 - 1.08) | 1.00<br>(0.94 - 1.06) |
| Gender of the child (Female vs. Male) | 1.01<br>(0.62 - 1.64) | 0.80<br>(0.50 - 1.30) | 1.05<br>(0.65 - 1.69) | 0.89<br>(0.57 - 1.38) | 0.96<br>(0.61 - 1.49) | 1.11<br>(0.71 - 1.74) | 0.90<br>(0.58 - 1.41) | 0.90<br>(0.56 - 1.46) |
| Age of the parent in years (linear) | 0.97<br>(0.93 - 1.01) | 1.00<br>(0.97 - 1.04) | 0.98<br>(0.94 - 1.02) | 0.99<br>(0.96 - 1.02) | 0.99<br>(0.95 - 1.02) | 0.99<br>(0.95 - 1.02) | 0.99<br>(0.96 - 1.03) | 0.99<br>(0.95 - 1.03) |
| Gender of the parent (Female vs. Male) | **0.53\***<br>**(0.31 - 0.88)** | 0.97<br>(0.58 - 1.63) | 1.28<br>(0.75 - 2.19) | 0.86<br>(0.53 - 1.39) | 0.99<br>(0.61 - 1.63) | 0.79<br>(0.49 - 1.28) | 0.83<br>(0.51 - 1.35) | 1.17<br>(0.69 - 1.97) |
| Parent level of education (Ref: At most High school graduate) |  |  |  |  |  |  |  |  |
| Some college | 0.94<br>(0.37 - 2.36) | 0.95<br>(0.35 - 2.56) | 0.89<br>(0.35 - 2.30) | 0.61<br>(0.25 - 1.46) | 0.80<br>(0.33 - 1.97) | 0.66<br>(0.27 - 1.61) | 1.72<br>(0.70 - 4.26) | 2.29<br>(0.89 - 5.90) |
| College graduate | 1.15<br>(0.48 - 2.79) | 1.36<br>(0.52 - 3.55) | 1.01<br>(0.41 - 2.50) | 0.99<br>(0.42 - 2.30) | 1.31<br>(0.55 - 3.13) | 0.74<br>(0.32 - 1.74) | 2.18<br>(0.90 - 5.28) | 1.28<br>(0.53 - 3.12) |
| Graduate degree | 0.49<br>(0.17 - 1.41) | 1.66<br>(0.57 - 4.82) | 0.92<br>(0.33 - 2.59) | 0.66<br>(0.25 - 1.70) | 0.79<br>(0.29 - 2.15) | 0.75<br>(0.28 - 1.97) | 2.62<br>(0.97 - 7.08) | 2.27<br>(0.81 - 6.34) |

\* p<0.05; \*\* p<0.01
1. Parental optimism represents the likelihood that a parent considers their child less likely or much less likely than average to develop the disease
2. Results were estimated using some imputation in order to overcome missing data (0-10% of data for all independent variables, see Appendix 2 for de

**Table 7a.**
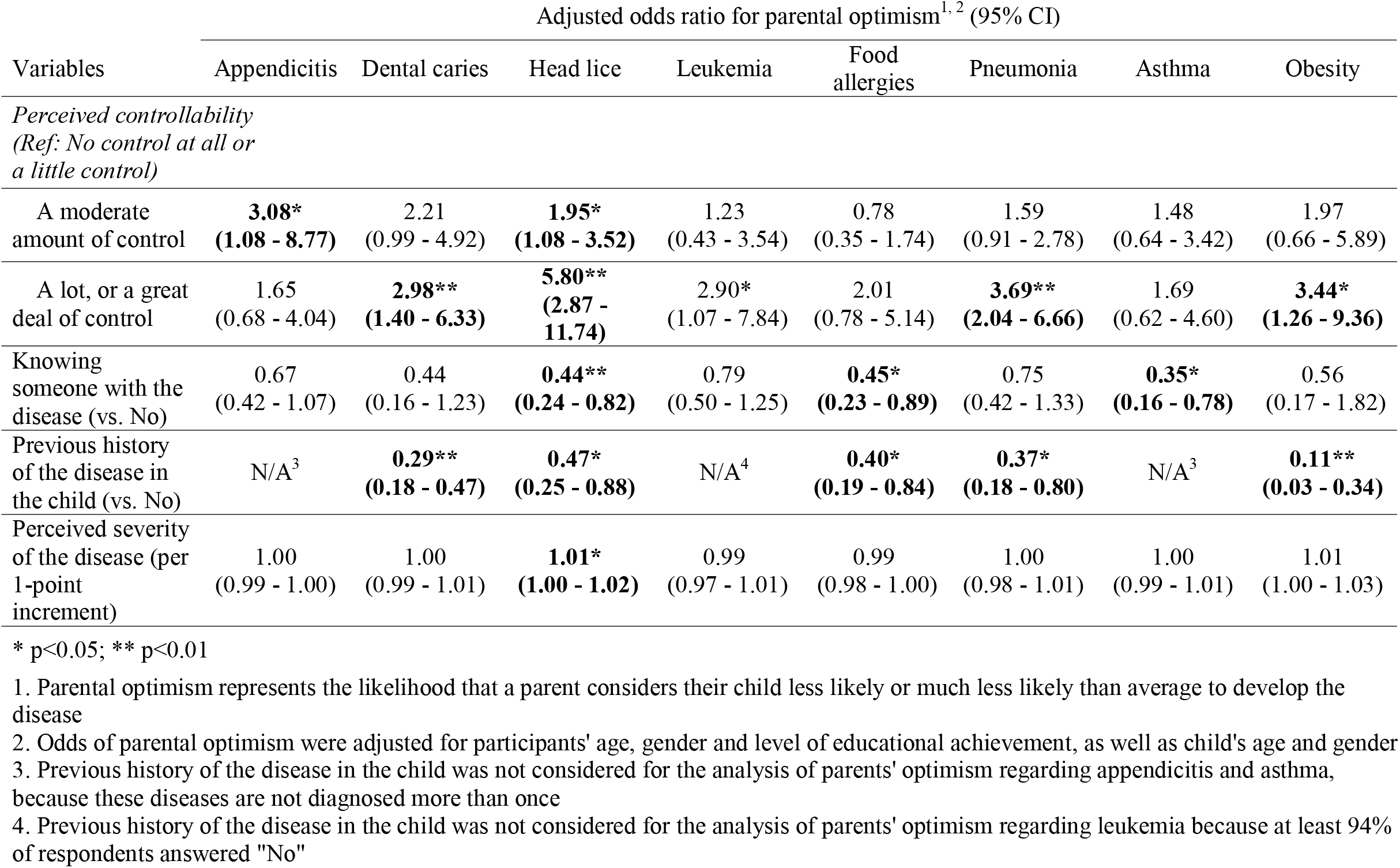
Sensitivity analysis using observed data without imputation: Odds of parental optimism regarding their child’s likelihood of developi experience and perceived disease severity.

| Variables | Adjusted odds ratio for parental optimism<sup>1,2</sup> (95% CI) |  |  |  |  |  |  |  |
| --- | --- | --- | --- | --- | --- | --- | --- | --- |
|  | Appendicitis | Dental caries | Head lice | Leukemia | Food allergies | Pneumonia | Asthma | Obesity |
| *Perceived controllability*<br>(Ref: No control at all or a little control) |  |  |  |  |  |  |  |  |
| A moderate amount of control | **3.08\***<br>(**1.08 - 8.77**) | 2.21<br>(0.99 - 4.92) | **1.95\***<br>(**1.08 - 3.52**) | 1.23<br>(0.43 - 3.54) | 0.78<br>(0.35 - 1.74) | 1.59<br>(0.91 - 2.78) | 1.48<br>(0.64 - 3.42) | 1.97<br>(0.66 - 5.89) |
| A lot, or a great deal of control | 1.65<br>(0.68 - 4.04) | **2.98\*\***<br>(**1.40 - 6.33**) | **5.80\*\***<br>(**2.87 - 11.74**) | 2.90*<br>(1.07 - 7.84) | 2.01<br>(0.78 - 5.14) | **3.69\*\***<br>(**2.04 - 6.66**) | 1.69<br>(0.62 - 4.60) | **3.44\***<br>(**1.26 - 9.36**) |
| Knowing someone with the disease (vs. No) | 0.67<br>(0.42 - 1.07) | 0.44<br>(0.16 - 1.23) | **0.44\*\***<br>(**0.24 - 0.82**) | 0.79<br>(0.50 - 1.25) | **0.45\***<br>(**0.23 - 0.89**) | 0.75<br>(0.42 - 1.33) | **0.35\***<br>(**0.16 - 0.78**) | 0.56<br>(0.17 - 1.82) |
| Previous history of the disease in the child (vs. No) | N/A<sup>3</sup> | **0.29\*\***<br>(**0.18 - 0.47**) | **0.47\***<br>(**0.25 - 0.88**) | N/A<sup>4</sup> | **0.40\***<br>(**0.19 - 0.84**) | **0.37\***<br>(**0.18 - 0.80**) | N/A<sup>3</sup> | **0.11\*\***<br>(**0.03 - 0.34**) |
| Perceived severity of the disease (per 1-point increment) | 1.00<br>(0.99 - 1.00) | 1.00<br>(0.99 - 1.01) | **1.01\***<br>(**1.00 - 1.02**) | 0.99<br>(0.97 - 1.01) | 0.99<br>(0.98 - 1.00) | 1.00<br>(0.98 - 1.01) | 1.00<br>(0.99 - 1.01) | 1.01<br>(1.00 - 1.03) |

\* p<0.05; \*\* p<0.01
1. Parental optimism represents the likelihood that a parent considers their child less likely or much less likely than average to develop the disease 2. Odds of parental optimism were adjusted for participants' age, gender and level of educational achievement, as well as child's age and gender 3. Previous history of the disease in the child was not considered for the analysis of parents' optimism regarding appendicitis and asthma, because these diseases are not diagnosed more than once 4. Previous history of the disease in the child was not considered for the analysis of parents' optimism regarding leukemia because at least 94% of respondents answered "No"

**Table 7b.** Sensitivity analysis using estimates by inverse probability weighting: Odds of parental optimism regarding their child’s likelihood of developing eight diseases, depending on parental perceived controllability, experience and perceived disease severity.

| Variables | Adjusted odds ratio for parental optimism<sup>1, 2</sup> (95% CI) |  |  |  |  |  |  |  |
| --- | --- | --- | --- | --- | --- | --- | --- | --- |
|  | Appendicitis | Dental caries | Head lice | Leukemia | Food allergies | Pneumonia | Asthma | Obesity |
| *Perceived controllability (Ref: No control at all or a little control)* |  |  |  |  |  |  |  |  |
| A moderate amount of control | **3.07\***<br>(**1.09 - 8.66**) | 2.33<br>(1.00 - 5.42) | **2.13\***<br>(**1.18 - 3.82**) | 1.20<br>(0.42 - 3.44) | 0.76<br>(0.32 - 1.78) | 1.59<br>(0.91 - 2.78) | 1.64<br>(0.70 - 3.83) | 2.34<br>(0.72 - 7.54) |
| A lot, or a great deal of control | 1.66<br>(0.61 - 4.53) | **3.29\*\***<br>(**1.47 - 7.34**) | **6.01\*\***<br>(**2.89 - 12.50**) | **2.71\***<br>(**1.00 - 7.29**) | 2.01<br>(0.82 - 4.92) | **3.69\*\***<br>(**2.04 - 6.66**) | 1.81<br>(0.68 - 3.83) | **3.77\***<br>(**1.26 - 11.25**) |
| Knowing someone with the disease (vs. No) | 0.63<br>(0.39 - 1.05) | 0.40<br>(0.15 - 1.12) | **0.46\***<br>(**0.25 - 0.87**) | 0.81<br>(0.52 - 1.28) | **0.45\***<br>(**0.23 - 0.88**) | 0.75<br>(0.42 - 1.33) | **0.31\*\***<br>(**0.13 - 0.72**) | 0.49<br>(0.13 - 1.81) |
| Previous history of the disease in the child (vs. No) | N/A<sup>3</sup> | **0.28\*\***<br>(**0.16 - 0.49**) | **0.47\***<br>(**0.25 - 0.88**) | N/A<sup>4</sup> | **0.43\***<br>(**0.20 - 0.92**) | **0.37\***<br>(**0.18 - 0.80**) | N/A<sup>3</sup> | **0.11\*\***<br>(**0.03 - 0.35**) |
| Perceived severity of the disease (per 1-point increment) | 0.99<br>(0.98 - 1.01) | 1.00<br>(0.99 - 1.01) | 1.01<br>(1.00 - 1.02) | 0.99<br>(0.97 - 1.01) | 0.99<br>(0.98 - 1.00) | 1.00<br>(0.98 - 1.01) | 1.00<br>(0.99 - 1.01) | 1.01<br>(1.00 - 1.03) |

\* p<0.05; \*\* p<0.01
1. Parental optimism represents the likelihood that a parent considers their child less likely or much less likely than average to develop the disease 2. Odds of parental optimism were adjusted for age, gender and level of educational achievement, as well as child's age and gender 3. Previous history of the disease in the child was not considered for the analysis of parents' optimism regarding appendicitis and asthma, because these diseases are not diagnosed more than once 4. Previous history of the disease in the child was not considered for the analysis of parents' optimism regarding leukemia because at least 94% of respondents answered "No"

## Discussion

In this study, a majority of parents displayed optimism when asked to estimate their child’s likelihood of developing eight different diseases compared to an average child of the same age. Furthermore, for most diseases, those who perceived greater controllability were more likely to display optimism. Conversely, parents with prior experience with a disease were less likely to exhibit optimism.

Optimism, measured in this way, refers to direct comparative optimism, which is elicited by inquiring about the risk of a given outcome (their child developing one of the eight diseases) compared to the average. This is in contrast with the indirect method, in which participants provide separate estimates of personal (in this case, their child’s) risk, and the risk estimate for a reference population. (Helweg-Larsen & Shepperd, 2001)

Our results are consistent with previous studies examining parental comparative optimism. One study reported high levels of parental optimism with regards to excessive weight gain in their children (86% of parents believed their child was less likely than average to gain excessive weight). (Drouin et al., 2019) Similarly, others have found parents to consider their child’s risk to become overweight or obese in the future as lower than for another “typical child in their community” (21% vs 72% risk). (Wright et al., 2017) A third study evaluated the perceived absolute risk difference of such estimates, showing that parents believed their child’s likelihood of becoming overweight to be 26.6% lower than that of a typical child at age 30. (Wright et al., 2018) Parental comparative optimism has also been reported regarding children’s risk of asthma exacerbation, (Shepperd et al., 2018) as well as tobacco, opioid and cannabis use. (Chadi et al., 2020; Drouin et al., 2019) To our knowledge, no previous study has examined the prevalence of parental comparative optimism regarding other pediatric disease.

Our findings are also consistent with existing literature on potential factors influencing comparative optimism in adults evaluating risks for themselves. Previous studies have shown a strong association between perceived controllability and comparative optimism in relation to various health outcomes. (Harris, 1996; Helweg-Larsen & Shepperd, 2001; Klein & Helweg-Larsen, 2002; Weinstein, 1980) Similarly, previous studies have also reported lower levels of comparative optimism regarding diseases with which people have had prior personal experience. (Helweg-Larsen & Shepperd, 2001; Weinstein, 1980) Third, optimism has generally been shown to be unaffected by perceived disease severity. (Helweg-Larsen & Shepperd, 2001; Weinstein, 1980; Weinstein, 1987) However, to our knowledge, no study to date has examined perceived controllability, prior personal experience or perceived disease severity as influencing *parental* comparative optimism regarding their child’s likelihood of disease.

A few limitations should be considered when interpreting the results of this study. First, data were obtained from a convenience sample recruited online, which *a priori* may not be representative of the general population. However, a number of reviews have validated Amazon’s MTurk as a reliable tool for conducting online behavioral and survey research, benefiting from a diverse pool of subjects. (Crump et al., 2013) In addition, our findings are also consistent with previous studies conducted with parents in clinical settings, (Drouin et al., 2019) further supporting the external validity of our results. Second, an inherent limitation of the methodology used in this study is that it only exposes unrealistic optimism at the group level. (Shepperd et al., 2013) Without further information on each child’s objective likelihood of disease, it is impossible to know whether any one participant’s optimism is truly unrealistically optimistic, or actually realistic or pessimistic. Nevertheless, it is noteworthy that parents displayed optimism regarding all diseases, including appendicitis and leukemia for which there are no known protective factors, and would therefore need to be considered unrealistic.

The results of this study are the first to suggest that perceived controllability and prior experience with a disease are factors also at play in parental *proxy* risk perception and resultant comparative optimism. These findings could have clinical and public health implications as they shed further light on this barrier to health-promoting behaviors in parents and their children. In a clinical setting, it may be helpful to understand parental perception of their child’s risk of a given condition or outcome in discussions about preventive measures, such as vaccinations, when discussing health behaviours, and when engaging in shared decision making. Eliciting information regarding parents’ perception of their child’s risk, as well as their perception of the controllability of certain diseases, may help healthcare professionals effectively identify parents displaying higher levels of optimism. This could be done prior to sharing risk information, and enable personalization of health-promoting messages. (Fowler & Geers, 2015) Furthermore, understanding factors underlying unrealistic optimism may help understand why people maintain optimism in the face of evidence that ought to eliminate it. For example, parents may be less attentive to novel risk information if they have never experienced the disease firsthand, and it may be helpful to focus on this particular element to mitigate unrealistic optimism (for example, by providing vivid descriptions or narratives of the disease first, in order to compensate for the lack of parental personal experience). (De Graaf et al., 2016) Although these clinical implications remain speculative, such insights may be taken into consideration in designing prevention interventions for conditions that may be the subject of parental unrealistic optimism.

## Conclusion

Overall, a majority of parents believed their child was less likely than average to develop each of eight selected diseases. Optimism was more likely if parents perceived a particular disease to be more controllable. Conversely, parents having prior experience with a disease were less likely to display optimism. Such insights may help develop effective interventions enhancing parental counseling and child health behaviors. Future research should examine other potential moderators (e.g., mood, perceived social stigma (Helweg-Larsen & Shepperd, 2001; Weinstein, 1987)) and include objective assessments of behavioral outcomes. Ideally these studies should employ prospective and/or interventional designs to better explore the interplay between unrealistic optimism, its moderators, various debiasing interventions and health behaviors.

## Data Availability

All data produced in the present study are available upon reasonable request to the authors

## Abbreviations

aOR: Adjusted odds ratio
CI: Confidence Interval
IQR: Interquartile Range
IPW: Inverse Probability Weighting
OR: Odds ratio

